# Fetal malnutrition and its predictors among term newborns in southern Ethiopia: a multicenter cross-sectional study

**DOI:** 10.64898/2026.06.24.26356480

**Authors:** Tomas Yeheyis, Melaku Haile Likka

**Author notes:** **Corresponding author**: Tomas Yeheyis, Po.Box 21 Hawassa, Ethiopia.

## Abstract

**Introduction:** Low and middle-income countries suffer from a high burden of undernutrition. Fetal malnutrition negatively impacts a newborn’s body composition, metabolism, and enzymatic processes, predisposing the newborn to malnutrition during childhood period. This study aims to assess the magnitude of fetal malnutrition and its predictors in southern Ethiopia.

**Methods:** A facility-based cross-sectional study was conducted among 423 pairs of mother and term newborn delivered from February 1-28, 2025, at five randomly selected public hospitals in southern Ethiopia. The Clinical Assessment of Nutrition (CAN) Score is used to assess fetal malnutrition. Logistic regression was employed to identify predictors of fetal malnutrition. Statistical significance of the association was declared at p < 0.05.

**Results:** Among 423 newborns included in the study, 60 (14.1%) had fetal malnutrition. Newborns born to women with a placental weight of 519 grams or less had ten times higher odds of fetal malnutrition compared to their counterparts (AOR=9.795, 95% CI: 4.881–19.657). Dietary counselling during pregnancy reduced odds of fetal malnutrition by 62.3% (AOR=0.377, 95% CI: 0.162–0.877); similarly, an extra meal during pregnancy was associated with reduced odds of fetal malnutrition by 71.6% (AOR=0.284, 95% CI: 0.131–0.616). Newborns delivered from women who had a MUAC (Mid Upper Arm Circumference) >22 cm had 75.7% lower odds of fetal malnutrition (AOR=0.243, 95% CI 0.074 –0.797), whereas maternal chronic medical illness increased the odds by threefold (AOR=3.419, 95% CI: 1.269–9.153).

**Conclusion:** There is a high magnitude of fetal malnutrition in the study area. Placental weight, dietary counselling, extra meals during pregnancy, MUAC and chronic medical illness were predictors of fetal malnutrition, signifying the need for a comprehensive approach targeting maternal nutrition during pregnancy.

## Introduction

Malnutrition is both a contributor to and a result of poverty and deprivation. Low and middle-income countries (LMICs) bear a disproportionately high burden of undernutrition [1]. Globally, 42.8 million children under five were wasted, and 150.2 million were stunted in 2024. Undernutrition, which primarily occurs in LMICs, is linked to nearly half of the deaths among children under five [1].

Fetal malnutrition (FM) is the intrauterine failure of the fetus to develop sufficient subcutaneous fat and muscle mass, regardless of birth weight or gestational age [2,3]. Although childhood undernutrition has received a lot of attention, fetal malnutrition is still not well addressed.

Fetal malnutrition has a wide span of short-term and long-term consequences [3–5]. During intrauterine life, it impacts neonatal body composition, metabolic function, and enzymatic processes, increasing the risk of hypoglycemia, hypothermia, and neurologic impairments [4,6]. Beyond the neonatal period, FM contributes to impaired physical growth, cognitive development, increased susceptibility to infection, and poorer educational outcomes later in life [5,7].

The burden of FM in LMICs, particularly in Sub-Saharan Africa (SSA), is likely higher in association with a higher magnitude of maternal undernutrition, infection, and limited access to maternal health care services. However, data on the magnitude and determinants of fetal malnutrition are limited compared to other types of undernutrition. Fetal malnutrition is often underdiagnosed due to the limited use of standardized assessment tools and reliance on birth weight alone.

Undernutrition is a serious public health problem in Ethiopia. Recent estimates indicate that only 11% of children aged 6–23 months receive the minimum acceptable diet, 37% of children under five years of age are stunted, 7% are wasted, and 21% are underweight [8]. Even if the government has put programs in place to improve the nutritional conditions of children and pregnant women, additional efforts are required to achieve national and international goals.

Fetal malnutrition has been linked to several maternal and pregnancy-related variables, including antenatal care (ANC) follow-up, maternal body mass index (BMI), iron-folic acid (IFA) provision, birth weight, placental weight, maternal infection during pregnancy, and antepartum hemorrhage. Nevertheless, the generalizability of these results is limited because they are primarily based on a single centre [6,9,10].

Fetal malnutrition is still poorly understood in Ethiopia, with no multi-centre data on its prevalence and determinants despite its clinical and public health significance. This study aimed to evaluate the magnitude of FM and identify its determinants in southern Ethiopia, and contribute to guiding interventions along the continuum of care from pregnancy to childhood

## Methods and materials

### Study area and design

An institution-based cross-sectional study was conducted at public hospitals of the Sidama Region from February 1–28, 2025. Sidama Regional State is one of the regional states in southern Ethiopia and was established in 2020 after its separation from the former Southern Nations, Nationalities and Peoples’ Regional State (SNNPR). The administrative capital of the region is Hawassa. The region has 19 public hospitals providing maternal and newborn health services to the population.

### Population

Randomly selected mothers who gave birth at the selected public hospitals of the Sidama region during the study period, along with their newborns, and who met the eligibility criteria.

### Eligibility criteria Inclusion criteria

All mothers who delivered a term single newborn baby at the randomly selected public hospitals during the study period were included in the study along with their newborns.

### Exclusion criteria

Mothers who were unable to participate in the study due to illness, births with an incomplete placenta, newborns with visible congenital abnormalities, preterm babies (<37 weeks of gestation), and newborns requiring immediate critical care were excluded.

### Sample size and sampling technique

Sample size was determined by using the single population proportion formula with a 95% confidence level, 5% margin of error, and a population proportion of 50. By accounting for a 10% non-response rate (i.e., divided by an expected response rate of 0.9), the final sample size came to be 427. From the 19 public hospitals available in the region during the study period, five (Hawassa University Comprehensive Specialized Hospital, Yirgalem General Hospital, Dore Bafana Primary Hospital, Leku General Hospital, and Adare General Hospital) were selected randomly. The sample was allotted proportionally to each hospital based on their delivery volume during the same period in the previous year. Systematic sampling techniques were employed to enrol participants. A sampling interval of 2 was used by dividing the expected number of women by the sample size to select participants, where the first respondent in each hospital was chosen randomly from the first two mothers (Fig 1. Sampling procedure among women who delivered at public hospitals of Sidama Region, Ethiopia, 2025)

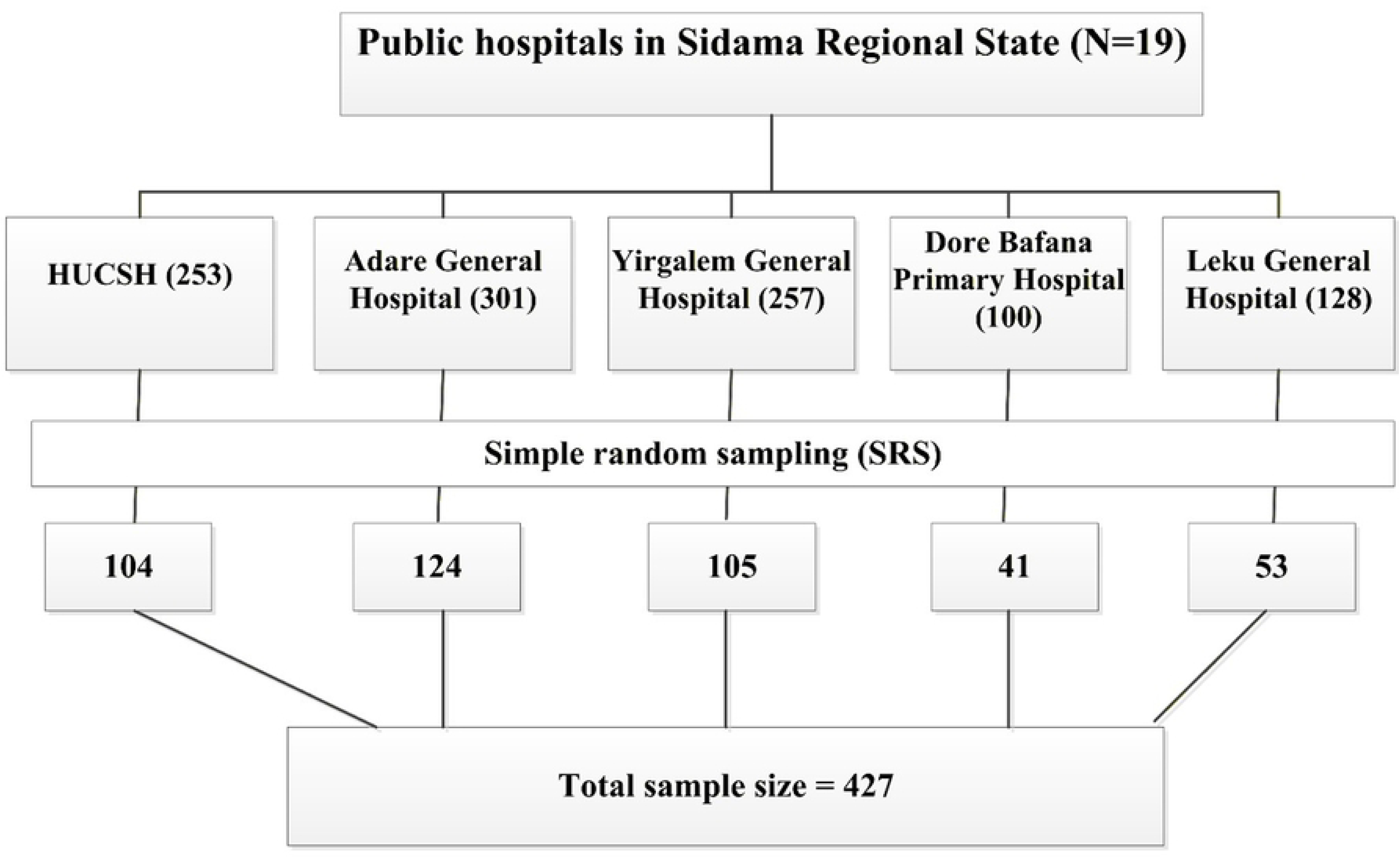

### Data collection procedure and operational definitions

Data were collected using a standardized nutrition assessment tool and a structured questionnaire adapted from previously published relevant literature [9–11]. Five experienced midwives, one from each hospital, collected data using KoBoToolbox electronically. Data collectors interviewed mothers once both the mothers and babies were stable after birth. MUAC was measured using a non-stretchable MUAC tape (DE20215134U1-Atelier Gardeur, Germany) at the midpoint between the acromion process of the shoulder and the olecranon process of the elbow of the left arm. Maternal undernutrition was defined as MUAC <22 cm[12]. Newborn’s birthweight and placental weight were measured using a calibrated digital baby scale (MS4200, Charder, Taiwan). All anthropometric measurements were recorded to the nearest 0.1 unit [13,14].

Fetal malnutrition was assessed within 24 hours of birth using the Clinical Assessment of Nutrition (CAN) score. The CAN score evaluates nine superficial and detectable signs of nutritional status (hair, cheek, chin and neck, arm, back, buttock, leg, chest, and abdomen of fetal malnutrition [15]. Each sign was scored from 1 to 4 based on inspection and physical examination of subcutaneous tissue stores, yielding a total score ranging from 9 to 36. Newborn babies with a CAN score <25 were classified as having fetal malnutrition[16]. In addition, data on sociodemographic characteristics of the women and newborns, obstetrics and medical-related characteristics, and nutritional behavior were collected through interviews and the client’s chart review.

### Data processing and analysis

Data were managed and analyzed using STATA version 19.5. A binary logistic regression model was applied to assess the statistical association between the outcome variable and each independent variable. Variables that showed statistical significance during bivariate analysis (p-value ≤ 0.25) were entered into the multivariate logistic regression method. The strength of associations was estimated using adjusted odds ratios (AOR) with 95% confidence intervals (CI), and statistical significance was determined at a p-value < 0.05.

### Data quality control

Training was provided to data collectors and supervisors over two days, and the data collection tool was pretested in 5% of the sample size at Melka Oda General Hospital. The weighing scale was calibrated before each measurement.

### Ethical Considerations

Ethical clearance was obtained from the Institutional Review Board of Hawassa University, College of Medicine and Health Sciences (IRB/421/17). Written informed consent was obtained from participants before their participation. The study was conducted in accordance with the Helsinki Declaration.

## Results

### Socio-demographic characteristics

A total of 423 mothers and their singleton newborns participated in the study (99% response rate). The mean + standard deviation (SD) age of the participating mothers was 26.1+ 4.7 years, with the plurality (40.5%) belonging to the 25-19 years age group. The majority of respondents were married (414 or 97.9%). More than one-third (35.9%) reported a monthly household income of 5,000-9,999 Ethiopian Birr (ETB). Regarding educational status, 31.6% had completed primary education. Over half of the participants (51.5%) were housewives, and 52.7% reported a family size of 4-6 members (Table 1).

**Table 1.** Socio-demographic characteristics of study participants.

| Variable | Frequency | Percent |
| --- | --- | --- |
| Age |  |  |
| 15-19 | 20 | 4.7 |
| 20-24 | 127 | 30.1 |
| 25-29 | 171 | 40.5 |
| 30-34 | 73 | 17.2 |
| 35-39 | 31 | 7.3 |
| 40-44 | 1 | 0.2 |
| Marital status |  |  |
| Single | 7 | 1.6 |
| Married | 414 | 97.9 |
| Divorced | 2 | 0.5 |
| Level of education |  |  |
| Cannot read and write | 72 | 17 |
| Primary education | 133 | 31.5 |
| Secondary education | 91 | 21.5 |
| Diploma | 55 | 13 |
| Degree | 67 | 15.8 |
| Masters/PhD | 5 | 1.2 |
| Occupation |  |  |
| Farmer | 18 | 4.3 |
| Government employee | 81 | 19.1 |
| Housewife | 218 | 51.5 |
| Merchant | 62 | 14.7 |
| Private employee | 13 | 3.1 |
| Other | 31 | 7.3 |
| Husband education |  |  |
| Cannot read and write | 54 | 12.8 |
| Primary education | 116 | 27.5 |
| Secondary education | 40 | 9.5 |
| Diploma | 26 | 6.2 |
| Degree | 85 | 20.2 |
| Masters/PhD | 100 | 23.8 |
| Husband occupation |  |  |
| Daily Laborer | 18 | 4.3 |
| Farmer | 73 | 17.3 |
| Government employee | 128 | 30.4 |
| Merchant | 86 | 20.4 |
| Private employee | 31 | 7.4 |
| Others | 85 | 20.2 |
| Monthly income in ETB |  |  |
| 10000-14999 | 69 | 16.3 |
| 15000-19999 | 52 | 12.3 |
| 5000-9999 | 152 | 35.9 |
| <5000 | 107 | 25.3 |
| >=20000 | 43 | 10.2 |
| Family size |  |  |
| ≤3 | 162 | 38.3 |
| 4-6 | 223 | 52.7 |
| 7 or more | 38 | 9 |

### Obstetrics-related characteristics

Among the mother study participants, 285 (67.4%) were multiparous. Most women (372 or 87.9%) had at least one ANC visit during the index pregnancy, and 206 (55.4%) of these had attended 4-6 ANC visits.

Pregnancy-related complications were reported by 50 (11.80%) of participants. Among these complications, preeclampsia accounted for 19 (38%). Chronic medical illnesses were present in 26 (6.1%) of women, with hypertension representing 53.9% of these reported conditions.

Additionally, 12.8% of participants were prescribed medications, and more than one-third (37.3%) experienced nausea and vomiting during pregnancy (Table 2).

**Table 2.** Obstetrics-related characteristics of women who participated in the study.

| Variable | Frequency | Percent |
| --- | --- | --- |
| Parity |  |  |
| Primipara | 138 | 32.6 |
| Multipara | 254 | 60.2 |
| Grand multipara (5 or more) | 31 | 7.2 |
| ANC follow-up |  |  |
| Yes | 372 | 87.9 |
| No | 51 | 12.1 |
| Number of ANC visits (n=372) |  |  |
| <=3 visits | 54 | 14.55 |
| 4-6 visit | 206 | 55.4 |
| >=7 visits | 112 | 30.1 |
| Complication during pregnancy |  |  |
| Yes | 50 | 11.8 |
| No | 373 | 88.2 |
| Type of complication during pregnancy (n=50) |  |  |
| APH | 11 | 22 |
| Anemia | 3 | 6 |
| Other | 3 | 6 |
| PROM | 7 | 14 |
| Preeclampsia | 19 | 38 |
| Gestational DM | 7 | 14 |
| Chronic medical illness |  |  |
| Yes | 26 | 6.1 |
| No | 397 | 93.9 |
| Type of chronic medical illness (n=26) |  |  |
| Hypertension | 14 | 53.9 |
| Diabetes Mellitus | 8 | 30.7 |
| Cardiac problems | 2 | 7.7 |
| Other | 2 | 7.7 |
| Taken prescribed medication during pregnancy |  |  |
| Yes | 54 | 12.8 |
| No | 369 | 87.2 |
| Nausea and vomiting during pregnancy |  |  |
| Yes | 158 | 37.3 |
| No | 265 | 62.7 |

### Nutritional characteristics

Regarding maternal nutrition, 275 (65%) of participants reported consuming additional meals during pregnancy, and 317 (74.9%) received dietary counselling. Consistent with this, 352 (83.2%) reported taking IFA supplementation; however, only 77(18.34%) used additional prenatal supplements. Overall, 401 (94.8%) of women had a MUAC >22 cm (Table 3).

**Table 3.** Nutrition-related characteristics of women who participated in the study.

| Variable | Frequency | Percent |
| --- | --- | --- |
| Received counselling on diet |  |  |
| Yes | 317 | 74.9 |
| No | 106 | 25.1 |
| Extra meal during pregnancy |  |  |
| Yes | 275 | 65 |
| No | 148 | 35 |
| Iron folic acid |  |  |
| Yes | 352 | 83.2 |
| No | 71 | 16.8 |
| Duration of IFA in months<br>(n=352) |  |  |
| 1 | 14 | 4 |
| 2 | 28 | 8 |
| 3 | 152 | 43.2 |
| 4 | 59 | 16.8 |
| 5 | 19 | 5.4 |
| 6 | 80 | 22.6 |
| Taken additional prenatal<br>supplements |  |  |
| Yes | 77 | 18.3 |
| No | 346 | 81.7 |
| The duration of prenatal<br>supplements taken in months<br>(n=77) |  |  |
| 1 | 9 | 11.7 |
| 2 | 11 | 14.3 |
| 3 | 31 | 41.6 |
| 5 | 3 | 3.9 |
| >=6 | 22 | 28.5 |
| MUAC |  |  |
| <22 | 22 | 5.2 |
| ≥22 | 401 | 94.8 |

### Characteristics of the newborn

Of the total newborns, 216 (51.1%) were male. Most infants (331 or 78.3%) had a normal birth weight. In terms of placental weight, 284 (67.1%) had a placental weight greater than 519g.

Overall, 60 (14.2%) of newborns had fetal malnutrition (Table 4).

**Table 4.**
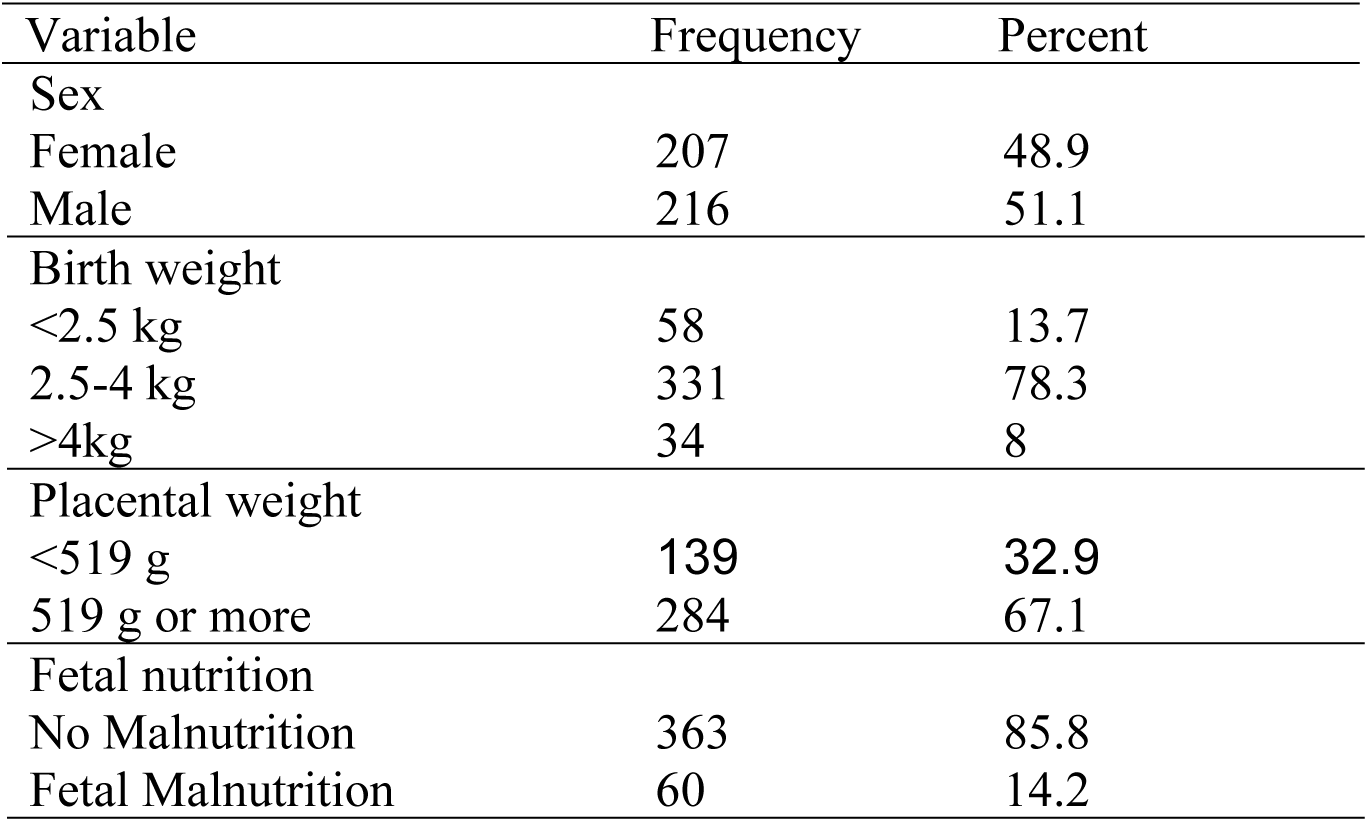
Fetal malnutrition and birth-related characteristics of newborns Variable Frequency Percent.

### Factors associated with fetal malnutrition

In the bivariable logistic regression analysis, placental weight, dietary counselling, extra meals during pregnancy, iron and folic acid supplementation, MUAC, and chronic medical illness were associated with fetal malnutrition at p< 0.25 and were included in the multivariable model. In the multivariable analysis, placental weight, dietary counselling, taking an extra meal during pregnancy, MUAC, and maternal chronic medical illness remained significantly associated with fetal malnutrition(p<0.05).

Newborns born to women with a placental weight of 519 g or less had more than nine times higher odds of fetal malnutrition compared to those with higher placental weight (AOR=9.795, 95% CI 4.881–19.657). Maternal receipt of dietary counselling during pregnancy lowers odds of fetal malnutrition by 62.3% (AOR=0.377, 95% CI: 0.162–0.877); similarly, an extra meal during pregnancy lowers the odds of fetal malnutrition by 71.6% (AOR=0.284, 95% CI: 0.131–0.616). On the other hand, newborns delivered from women who had a MUAC> 22 cm had 75.7% lower odds of fetal malnutrition compared to their counterparts (AOR=0.243, 95% CI: 0.074–0.797), whereas maternal chronic medical illness increased the odds by threefold AOR=3.419, 95% CI: 1.269 – 9.153) (Table 5)

**Table 5.**
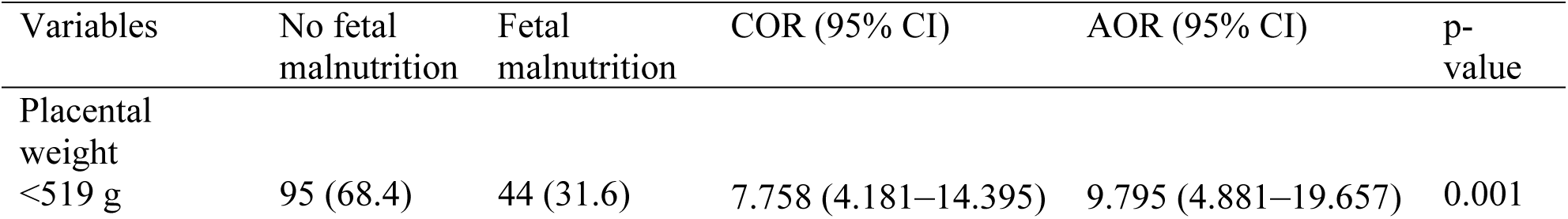
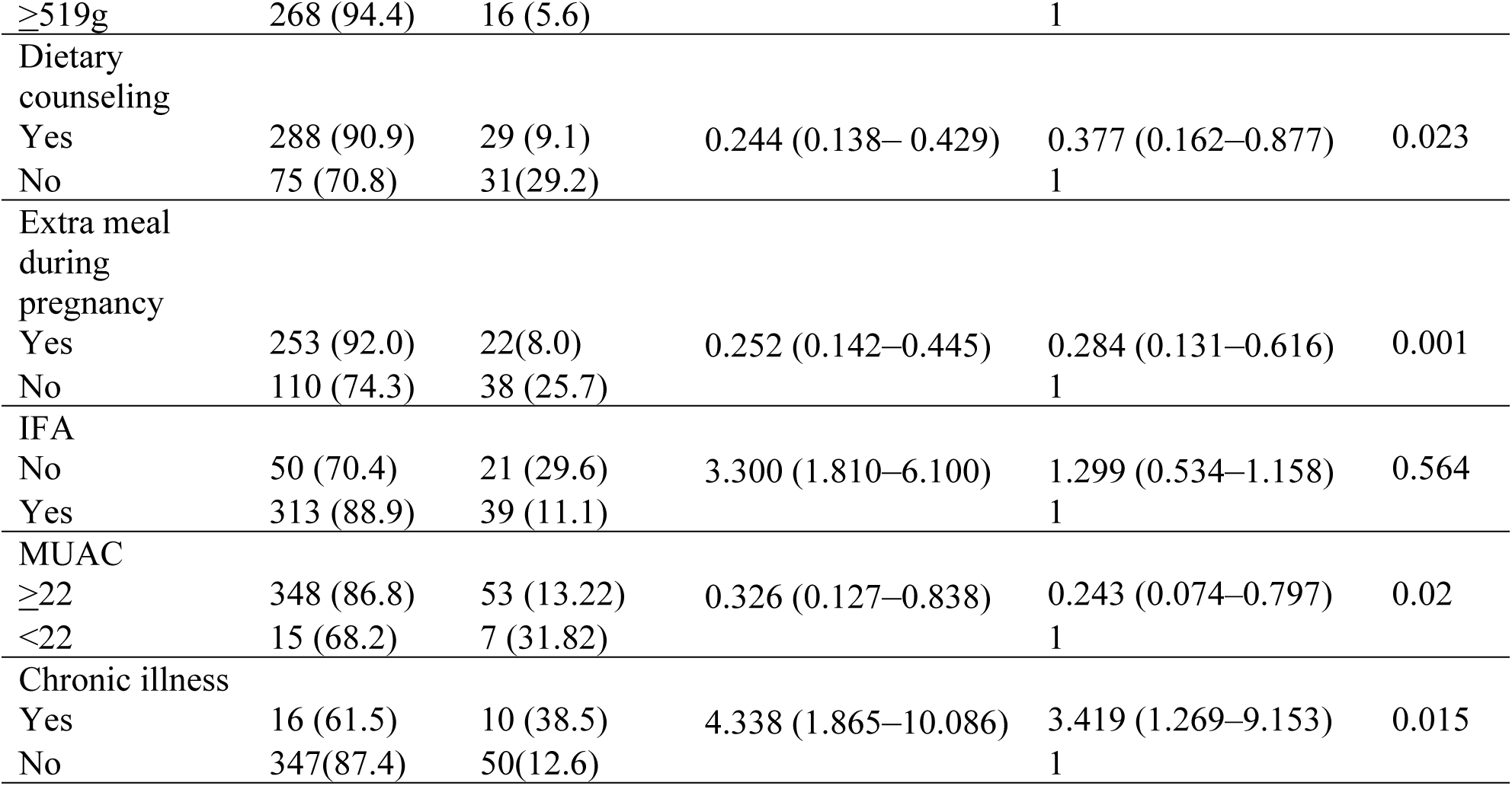
Factors associated with fetal malnutrition (n=423).

| Variables | No fetal malnutrition | Fetal malnutrition | COR (95% CI) | AOR (95% CI) | p-value |
| --- | --- | --- | --- | --- | --- |
| Placental weight |  |  |  |  |  |
| <519 g | 95 (68.4) | 44 (31.6) | 7.758 (4.181–14.395) | 9.795 (4.881–19.657) | 0.001 |
| ≥519g | 268 (94.4) | 16 (5.6) |  | 1 |  |
| Dietary counseling |  |  |  |  |  |
| Yes | 288 (90.9) | 29 (9.1) | 0.244 (0.138–0.429) | 0.377 (0.162–0.877) | 0.023 |
| No | 75 (70.8) | 31(29.2) |  | 1 |  |
| Extra meal during pregnancy |  |  |  |  |  |
| Yes | 253 (92.0) | 22(8.0) | 0.252 (0.142–0.445) | 0.284 (0.131–0.616) | 0.001 |
| No | 110 (74.3) | 38 (25.7) |  | 1 |  |
| IFA |  |  |  |  |  |
| No | 50 (70.4) | 21 (29.6) | 3.300 (1.810–6.100) | 1.299 (0.534–1.158) | 0.564 |
| Yes | 313 (88.9) | 39 (11.1) |  | 1 |  |
| MUAC |  |  |  |  |  |
| ≥22 | 348 (86.8) | 53 (13.22) | 0.326 (0.127–0.838) | 0.243 (0.074–0.797) | 0.02 |
| <22 | 15 (68.2) | 7 (31.82) |  | 1 |  |
| Chronic illness |  |  |  |  |  |
| Yes | 16 (61.5) | 10 (38.5) | 4.338 (1.865–10.086) | 3.419 (1.269–9.153) | 0.015 |
| No | 347(87.4) | 50(12.6) |  | 1 |  |

## Discussion

This study has examined the magnitude of FM and its predictors in southern Ethiopia. The study has shown that 14.1% of newborns had fetal malnutrition. Placental weight, dietary counselling, taking extra meals during pregnancy, MUAC, and chronic medical illness during pregnancy were predictors of fetal malnutrition.

Even though the magnitude of FM observed in this study is in line with findings from previous studies conducted in northeast Ethiopia[10,17]. The finding shows that the burden of FM in the area is high, requiring attention. The magnitude observed in the study is lower than the findings of studies conducted in Ethiopia, Nigeria, and India [9,18,19,6,20]. The discrepancy in the findings is attributed to the differences in the study setting and population.

Newborns born to women with a placental weight of 519g or less had nearly ten times more fetal malnutrition compared to those with a placental weight of more than 519g. A similar finding is observed in studies conducted in Ethiopia and Nigeria [10,11,21]. The placenta is the primary line for the exchange of nutrients and oxygen to the fetus. The association between low placental weight and fetal malnutrition shows the significance of placental function for fetal growth and nutritional transfer. Reduced placental mass is often an indication of insufficient or poor placental development. This, in turn, limits the supply of nutrients required for fetal tissue deposition, which leads to inadequate subcutaneous fat and muscle mass. This is in line with the broader evidence available that links placental insufficiency with poor neonatal outcomes [18,22,23].

Maternal receipt of dietary counselling during pregnancy was associated with 77% lower odds of fetal malnutrition, compared to women who didn’t receive dietary counselling. This finding is consistent with the findings of a previous study conducted in northwest Ethiopia[10]. The significance of maternal nutrition education during pregnancy as a modifiable predictor of fetal health is indicated by the preventive impact of dietary counselling against fetal malnutrition.

Dietary counselling gives pregnant women the opportunity to improve their awareness of the nutrition needed during pregnancy and adherence to recommended nutritional protocols during pregnancy [24–26]. The study implies that nutritional education during antenatal pregnancy reduces the odds of occurrence of fetal malnutrition, highlighting the need to give emphasis to nutritional education during ANC.

The study highlights the impact of direct nutritional support during pregnancy. We have found that taking extra meals during pregnancy has significantly reduced the odds of fetal malnutrition (AOR=0.284, 95% CI 0.131-0.616). This finding is particularly resonant in the context of low-income countries, where maternal diets frequently fail to meet the heightened metabolic needs of pregnancy [27,28]. This finding is supported by evidence showing improved energy intake and dietary diversity with healthier birth weight, indicating a clear opportunity for intervention.

Mother’s baseline nutritional status is an equally important predictor of fetal malnutrition. A maternal MUAC greater than 22 was associated with a 75.7% drop in the odds of fetal malnutrition (AOR=0.243, 95% CI 0.074 – 0.797). This finding reinforces the clinical utility of MUAC as a proxy for maternal energy stores that are essential for preventing fetal growth restriction and buffering against nutritional deficiencies [29,30].

Maternal chronic medical illness increased the odds of fetal malnutrition nearly fourfold compared to their counterparts. The association observed between chronic medical illness and fetal malnutrition indicates the complex interplay between the mother’s health and the nutritional maturity of the fetus. Chronic medical illnesses like hypertension, diabetes, anemia, and infection alter metabolic processes, increase the body’s nutritional demand, reduce blood flow to the uterus and placenta, and in turn restrict the fetus’s access to nutrients. These pathophysiological alterations can impact the buildup of fat and lean body mass and interfere with normal embryonic growth patterns [17,31,32]. This finding emphasizes the need for early detection and treatment of maternal comorbidities within antenatal care since increasing mother health is critical to improving fetal nutritional outcomes and breaking the intergenerational cycle of malnutrition.

The high prevalence of fetal malnutrition in the study signifies a substantial burden of intrauterine nutritional deficiency. The findings underscore the need to strengthen ANC with a focus on early identification of high-risk pregnancies in relation to placental insufficiency and fetal low birth weight, emphasising nutrition education and management of underlying conditions.

The study has several strengths, including the use of a standard CAN score method to assess the nutritional status of a newborn, employing the study in multiple facilities of different levels, ranging from primary hospitals to a referral centre, which increases the likelihood of facing women of different socio-cultural backgrounds and health levels. But the study is not free of limitations, which include the fact that it was conducted only among women delivered in public hospitals and does not include home delivery, which can undermine the true magnitude of the problem in the study setting. Furthermore, since the assessment of fetal malnutrition is based on physical examination, there could be a risk of misclassification.

### Conclusion

The study has found a high burden of fetal malnutrition in the study area. Fetal malnutrition was significantly associated with placental weight, nutritional counselling, taking extra meals during pregnancy, maternal MUAC, and the presence of chronic medical illnesses. The finding highlights the multifactorial nature of fetal malnutrition and the need for a comprehensive approach targeting maternal health and nutrition during pregnancy.

## Abbreviations

ANC, antenatal care; BMI, body mass index; CAN, Clinical Assessment of Nutrition; ETB, Ethiopian Birr; FM, fetal malnutrition; IFA, iron-folic acid; LMICs, low- and middle-income countries; MUAC, mid-upper arm circumference; SD, standard deviation; SNNPR, Southern Nations, Nationalities, and Peoples’ Regional State; SSA, Sub-Saharan Africa.

## Data Availability

Data is submitted to the system along with the manuscript

## Acknowledgement

We would like to acknowledge the Royal Society of Tropical Medicine and Hygiene for funding the project through the Early Career Researchers’ Grant. We are also grateful to the participants and data collectors. We would like to declare that we have used Grammarly (Free AI writing assistance) to correct grammar and for overall English writing assistance.

## Contribution of authors

TY conceptualized the study, did the analysis, and drafted the manuscript. MHL guided the analysis, reviewed the manuscript, and supervised the study process.

